# Content Analysis of YouTube Videos Related to E-cigarettes and COVID-19

**DOI:** 10.1101/2023.01.06.23284266

**Authors:** Juhan Lee, Dhiraj Murthy, Grace Kong

## Abstract

**INTRODUCTION:** E-cigarettes are frequently promoted on social media and portrayed in ways that are attractive to youth. While COVID-19 pandemic significantly affected people’s lives, less known is how the pandemic influenced e-cigarette-related marketing and information on social media. This study identifies how e-cigarettes are portrayed during the COVID-19 pandemic on YouTube, one of the most popular social media platforms.

**METHODS:** We searched for combinations of search terms related to e-cigarettes (i.e., “electronic cigarette”, “e-cigarette”, “e-cig”, “vape” and “vaping”) and COVID-19 (i.e., “corona”, “COVID”, “lockdown” and “pandemic”). To be included in the analysis, the video must be: uploaded after February 1, 2020, in English, related to e-cigarettes and COVID-19 and less than 30 minutes in length. We assessed video themes related to e-cigarettes and COVID-19, uploader characteristics, and featured e-cigarette products.

**RESULTS:** We examined N=307 videos and found that N=220 (73.6%) were related to the health effects of e-cigarette use on COVID-19, followed by videos of how COVID-19 affects e-cigarette access/sales (N=40, 12.9%), and face mask-related videos (N=16, 5.1%) which included content regarding masks and e-cigarette use. Instructional videos on how to modify e-cigarettes to use with masks had the highest number of likes (Median=23; IQR=32) and comments (Median=10; IQR=7).

**CONCLUSIONS:** This study identified various e-cigarette contents on YouTube during the COVID-19 pandemic. Our findings support the need for continuous surveillance on novel vaping-related content in reaction to policies and events such as the global pandemic on social media is needed.

## 1. INTRODUCTION

E-cigarette use exposes youth to nicotine and other toxicants, which is associated with nicotine addiction and progression to other tobacco product use.(1) Nevertheless, e-cigarette use is frequently portrayed as glamourous on social media, which may appeal to young people.(2,3) Manufacturers and retailers also use social media to market and sell e-cigarettes.(4) For example, YouTube, one of the social media platforms most frequently used by young people (95% among youth who aged 13-17 in 2022),(5) makes youth-appealing content such as vape tricks highly accessible. Concerningly, 48% of such video content has been posted by the tobacco industry accounts.(3,6) Given high social media use by youth and rapidly evolving e-cigarette promotional content on social media, understanding how e-cigarettes are portrayed on social media is urgently needed.

The COVID-19 pandemic affected all aspects of people’s lives across the globe and continues to do so. There is evidence that during the height of the COVID-19 pandemic, social media was home to misleading content espousing therapeutic and preventive effects of nicotine on COVID-19.(7) COVID-19 also appeared in the marketing content of e-cigarettes on social media.(8) For example, Puff Bar posted photos of its factory workers in China wearing masks and protective gear to demonstrate ‘health reassurances’ on Instagram.(8) One notable change in the marketing environment with shop closures during the COVID-19 pandemic as tobacco stores and vape shops were considered non-essential businesses. As such, some vape shops posted social media offering ‘contactless delivery’ and ‘curbside pickup’. Some posts included health assurance themes (e.g., claiming health benefits from e-cigarette use during COVID-19) and encouraging stockpiling of e-cigarette products during the pandemic.(8) This shift to online retailers is concerning because they generally have less restrictive age verification, which can lead to underage e-cigarette use.(9–11)

There is a dearth of work examining how COVID-19 is used in e-cigarette marketing on social media. YouTube is an important source to understand e-cigarette promotion. Given that YouTube is the most frequently used social media by all age groups,(5) and YouTube is a popular source to obtain health-related information, including e-cigarettes,(12) examining how COVID-19 and e-cigarettes are portrayed on YouTube is important. This information may provide valuable insight into prohibiting the marketing content that promotes e-cigarette use and identify misinformation to inform countermarketing and educational efforts. Understanding how e-cigarettes are portrayed in the context of COVID-19 on YouTube is important as this content may influence perception, attitudes, and even e-cigarette-related behaviors.(13,14)

This study developed a data collection and coding framework to identify and evaluate how COVID-19 and e-cigarettes are portrayed in YouTube videos. Among the videos that included e-cigarettes and COVID-19, we further identified the video’s themes, who posted these videos, what e-cigarette products were featured, and whether levels of engagement (e.g., number of views, comments, likes) differs by these video characteristics.

## 2. METHODS

### 2.1. Data Collection

We collected videos by combining search terms related to e-cigarettes and COVID-19 in July 2021 on YouTube. We limited our search to videos uploaded after February 1, 2020, when COVID-19 was widespread globally. E-cigarette-related terms included “electronic cigarette”, “e-cigarette”, “e-cig”, “vape” and “vaping” and COVID-related terms included “corona”, “COVID”, “lockdown” and “pandemic”. We included e-cigarette search words that have been shown to have the most connections to e-cigarette content on YouTube based on our previous study.(15) To be included in our dataset, the video had to: (1) be in English, (2) have content related to e-cigarettes and COVID-19, and (3) be less than 30 minutes in length (to ensure that we are capturing videos that people may watch). We also obtained video metadata (i.e., view counts, likes, dislikes, and upload date).

### 2.2. Codebook Development

We adapted a codebook that had previously been used for the analysis of e-cigarette-related content on YouTube for this study (**Table 1**).(12) The lead author viewed the videos and categorized themes (i.e., classified the major theme as it relates to e-cigarettes and COVID-19), uploader type (who posted these videos), and featured e-cigarette products (whether these videos portrayed e-cigarette products) based on prior research(3,12) and added new themes related to COVID-19. Another author reviewed and confirmed and/or modified the themes. We determined interrater reliability by coding approximately 10% of videos (N=40 videos). Gwet AC1 statistics,(16) which addressed the marginal predicted probability issues of Cohen’s kappa statistics,(17) ranged from 0.8558 to 1, which indicates “almost perfect” agreement.(18)

**Table 1.**
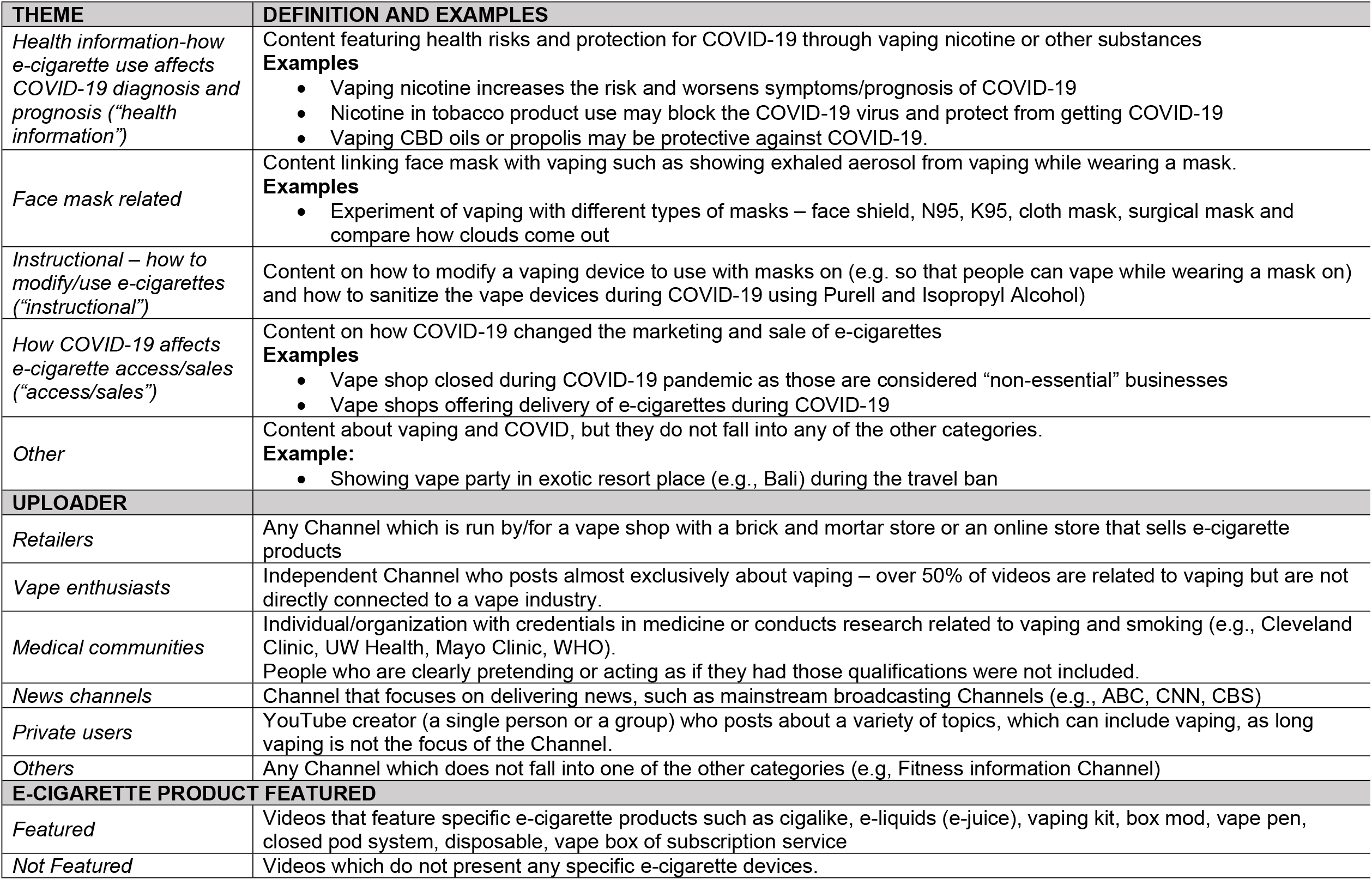
Definitions and examples of categories 16

### 2.3. Data Analysis

We conducted descriptive statistical analyses of the number of views, comments, likes, and upload years for each video theme, uploader type and featured e-cigarette product. Due to the non-normality of our data, we calculated the median and the inter-quartile range (IQR) for continuous variables (i.e., view, comment, like) and count and percentage for a categorical variable (i.e., upload year). To assess whether themes, uploader types and featured e-cigarette products had different levels of engagement (i.e., number of views, comments, likes) and upload year, we conducted the Kruskal-Wallis test for continuous variables and Pearson chi-square test for categorical variables. To assess whether video themes differed by uploader type, we also conducted chi-square tests.

## 3. RESULTS

Figure 1. illustrates our selection of e-cigarette- and COVID-19-related YouTube videos and exclusion rules. We obtained N=3,796 videos using combinations of e-cigarette-related and COVID-19-related terms (as described *2*.*1. Data Collection*) and included N=307 unique videos as final analytic sample per our inclusion criteria.

**Figure 1.**
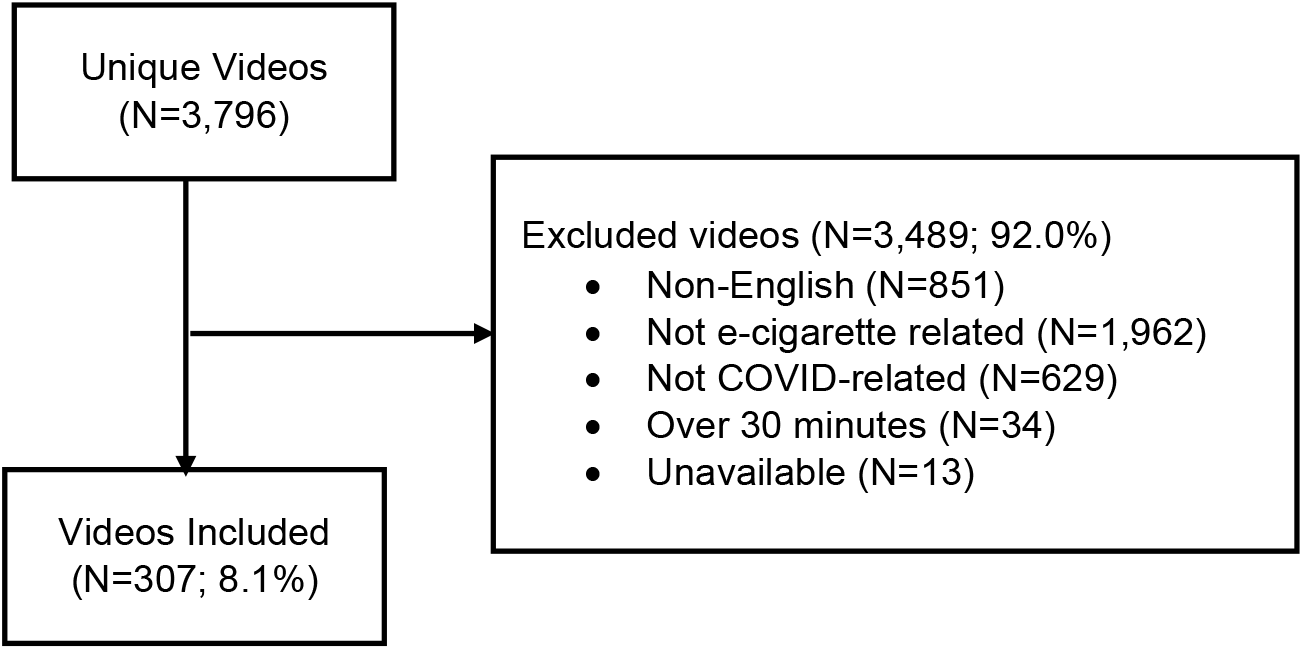
Inclusion of YouTube videos on vaping and COVID-19 and reasons for exclusion.

### 3.1. Video themes

There are five themes related to both e-cigarettes and COVID-19: (1) *health information regarding e-cigarettes and COVID-19* (“*health information*”); these videos portrayed whether vaping nicotine or other substances (e.g., CBD oils, propolis) increased the health risks related to COVID-19 or discussed potential therapeutic effects of vaping on COVID-19. (2) *Face mask-related* videos included content that links masks with vaping (“face masking”); these videos portrayed vaping while wearing a mask (e.g., showing and comparing visible vape clouds through different masks - KN95, N95, cloth masks, and surgical masks. (3) *Instructional – how to modify/use e-cigarettes* (“*instructional*”) videos; these videos portrayed how to content, including sanitizing vape devices during COVID-19 and demonstrated how to modify a vaping device to use with face masks. (4) *how COVID-19 affects e-cigarette access/sales* (“*access/sales*”) videos discussed how the COVID-19 pandemic changed the marketing and sale of e-cigarettes. This included the vape shop closing and offering delivery of vape products. (5) “*Other”* themes included videos that did not fit into these themes. These included videos of vape parties abroad during the travel ban.

### 3.2. Uploader type

To determine the uploader type, we examined the Channel Pages (i.e., “profile” page of the video uploader) of each YouTube video. There are six uploader types: (1) *Retailers* are any Channel that is run by/for vape and other tobacco brick and mortar stores or online stores where they sell e-cigarettes. (2) *Vape enthusiasts* are independent users who post almost exclusively about vaping – over 50% of their videos are related to vaping but are not directly connected to the vape industry. (3) *Medical communities* are any individual/organization with medical/healthcare credentials or who/that conducts research related to vaping and smoking (4) *News Channels* are media outlets that focus on delivering news, such as traditional broadcasting as well as new media channels. (5) *Private users* are YouTube creators (a single person or a group) who post about a variety of topics, which can include vaping. (6) *Other* include other Channels that do not fall into these categories such as a Channel focused on fitness.

### 3.3. Featured e-cigarette products

We determined first whether an e-cigarette product was present, and if present, we attempted to identify the depicted product. (1) “*Featured”:* when the video explicitly introduced or presented an e-cigarette product including any types of e-cigarette device including cigalike, e-liquid (e-juice), vaping kit, box mod, vape pen, closed pod system, disposable, or vape box of subscription service. (2) *“Not featured”:* when videos did not feature a specific e-cigarette product.

**Table 2** provides details of our results, including the number of views, comments, likes, and published year for each of the video themes, uploader types, and featured e-cigarette products. Among N=307 videos, 73.6% were “*health information*” videos (e.g., how e-cigarette use affects COVID-19 diagnosis and prognosis), followed by “*access/sales*” and “*face mask-related*” videos (e.g., experiment of vaping with different types of masks; 5.1%). 36.5% were posted by *private users*, followed by *news channels* (36.2%), *medical communities* (16.9%), and *retailers* (6.8%). Of note, the majority (89.3%) of the videos did not feature a specific e-cigarette product.

**Table 2.** Descriptive Characteristics of Videos Related to E-cigarettes and COVID-19 (N=307)

|  | Overall | 1. View <sup>a</sup> | 2. Comment <sup>a</sup> | 3. Like <sup>a</sup> | 4. year <sup>b</sup> |  |
| --- | --- | --- | --- | --- | --- | --- |
|  |  |  |  |  | 2020 | 2021 |
|  | N (%) | Median (IQR) | Median (IQR) | Median (IQR) | N (%) |  |
| Theme <sup>e</sup> |  | *p*=0.179 <sup>c</sup> | *p*=0.044 <sup>c</sup> | *p*=0.009 <sup>c</sup> | *p*=0.099 |  |
| Health information-how e-cigarette use affects COVID-19 diagnosis and prognosis (“*health information*”) | 229 (73.6) | 219 (1625) | 2 (8) | 3 (19) | 208 (76.7) | 21 (58.3) |
| Face mask related | 16 (5.1) | 41.5 (835.5) | 0 (1) | 2.5 (8) | 13 (4.8) | 3 (8.3) |
| Instructional – how to modify/use e-cigarettes (“*instructional*”) | 10 (3.2) | 392 (1646) | 10 (7) | 23 (32) | 6 (2.2) | 3 (8.3) |
| How COVID-19 affects e-cigarette access/sales (“*access/sales*”) | 40 (12.9) | 680 (3698) | 7 (36) | 22 (76) | 32 (11.8) | 7 (19.4) |
| Other | 16 (5.1) | 146.5 (2670) | 2.5 (6) | 4 (54) | 12 (4.4) | 2 (5.6) |
| Uploader |  | *p*=0.001 <sup>c</sup> | *p*=0.054 <sup>c</sup> | *p*=0.609 <sup>c</sup> | *p*=0.420 |  |
| Retailers | 21 (6.8) | 520 (1519) | 1 (7) | 12 (29) | 19 (7.0) | 2 (5.6) |
| Vape enthusiasts | 9 (2.9) | 48 (336) | 1 (6) | 3 (24) | 8 (3.0) | 1 (2.8) |
| Medical communities | 52 (16.9) | 457 (7229.5) | 1.5 (13.5) | 5.5 (47) | 47 (17.3) | 5 (13.9) |
| News Channels | 111 (36.2) | 319 (2003) | 3 (15) | 3 (20) | 102 (37.6) | 9 (25.0) |
| Private users | 112 (36.5) | 101 (737) | 1 (9.5) | 3 (27) | 93 (34.3) | 19 (52.8) |
| Others | 2 (0.7) | 213 (382) | 0 (0) | 2 (4) | 2 (0.7) | 0 (0) |
| E-cigarette product featured |  | *p*=0.697 <sup>c</sup> | *p*=0.718 <sup>c</sup> | *p*=0.547 <sup>c</sup> | *p*=0.365 |  |
| Featured | 33 (10.7) | 197 (1643) | 1 (11) | 3 (21) | 28 (10.3) | 2 (5.6) |
| Not Featured | 277 (89.3) | 230 (2013) | 2 (10) | 4 (28) | 243 (89.7) | 34 (94.4) |

a: Median (IQR) for continuous variables due to non-normality; b: N(%) indicated for categorical variables; c: Kruskal-Wallis test for continuous variables due to non-normality; d: Pearson chi-square tests for categorical variables; e: Themes are coded multiple times if the video presented different themes

There were significant differences between the video theme and the number of comments and likes. “*Instructional*” videos had the highest like count (Median=23; IQR=32), followed by “*access/sales*” videos (Median=22; IQR=76). Similarly, “*instructional*” videos had the highest number of comments (Median=10; IQR=7), followed by “*access/sales*” videos (Median=7; IQR=36). There was also a significant difference in view count by uploader type. Videos uploaded by *retailers* had the highest view count (Median=520; IQR=1519), followed by *medical communities* (Median=457; IQR=7229.5), and *news channels* (Median=319; IQR=2003). There was no significant difference in view counts, comments, likes, and upload year by featured e-cigarette product (all *ps*>0.05).

**Table 3** presented results related to video themes and uploader type. Importantly, we found that there was a significant association between theme and uploader type. Specifically, the majority of “*health information*” videos were uploaded by *medical communities* and *news channels*, 98.1% and 90.1%, respectively. The proportions of videos that were uploaded by *private users* were “*health information”* (59.8%), followed will by “*access/sales*” (16.1%) and “*face mask related*” (13.4%). The association between video themes posted by *retailers* and *vape enthusiasts* differed. For example, the most common videos that were uploaded by *retailers* were “*access/sales*” (47.6%) and “*instructional*” (19.1%); while “*health information”* (44.4%) and “*access/sales”* (22.2%) were most common videos that uploaded by *vape enthusiasts*.

**TABLE 3.** Proportions across themes and uploader types

|  | Uploader |  |  |  |  |  |  |
| --- | --- | --- | --- | --- | --- | --- | --- |
|  | Retailers | Vape enthusiasts | Medical communities | News Channels | Private users | Others |  |
|  | N (%) | N (%) | N (%) | N (%) | N (%) | N (%) | P-value |
| Theme |  |  |  |  |  |  | **<0.001** |
| Health information-how e-cigarette use affects COVID-19 diagnosis and prognosis (*"health information"*) | 5 (23.8) | 4 (44.4) | 51 (98.1) | 100 (90.1) | 67 (59.8) | 2 (100) |  |
| Face mask related | 0 (0) | 1 (11.1) | 0 (0) | 0 (0) | 15 (13.4) | 0 (0) |  |
| Instructional – how to modify/use e-cigarettes (*"instructional"*) | 4 (19.1) | 0 (0) | 0 (0) | 0 (0) | 5 (4.5) | 0 (0) |  |
| How COVID-19 affects e-cigarette access/sales (*"access/sales"*) | 10 (47.6) | 2 (22.2) | 0 (0) | 9 (8.1) | 18 (16.1) | 0 (0) |  |
| Other | 2 (9.5) | 2 (22.2) | 1 (1.9) | 2 (1.8) | 7 (6.3) | 0 (0) |  |

## 4. DISCUSSION

To the best of our knowledge, this study is the first to examine YouTube videos that are relared to both e-cigarettes and COVID-19. Moreover, our data represents the height of the pandemic, between February 2020 and July 2021. In our collected data, the most common theme was “*health information*”, which presented conflicting health information on whether vaping exacerbated the risk of or was protective from COVID-19 infection. The second most common theme was related to “*access/sales*”, which included the closure of vape shops and the shift to online stores due to the pandemic. We also observed that videos uploaded by retail stores showed the highest view counts, and “*instructional*” and “*access/sales*” videos showed the highest comments and likes, respectively.

The “*health information” videos* reflected media coverage of research studies published at the time, which conflicted over findings regarding the role of e-cigarette use on COVID-19. Some studies indicated a greater risk of COVID-19 infection and worse prognosis(19–22) since exposure to nicotine and other chemicals through e-cigarette use may affect the respiratory system.(23–28) In contrast, other studies indicated that nicotine and e-cigarette use may not be associated with COVID-19; rather, nicotine may even be a protective factor.(29–33) Only a few videos, typically uploaded by e-cigarette brand and retailers, either suggested that vaping is protective against COVID-19 or claimed no association. For example, one video uploaded by an online e-cigarette retailer claimed that *“NIDA did not provide scientific evidence when NIDA had advised quit vaping during pandemic*.” Of note, this video mimicked a news channel, including hosts wearing suits and text scrolling at the bottom of the screen, which may have misled viewers. This online retailer uploaded a follow-up video that claimed that FDA found no connection between vaping and getting COVID-19. This video did not provide valid sources and provided only a link to a pro-vaping organization blog with broken news links. A few videos recommended vaping other substances (e.g., CBD, propolis) to protect themselves from COVID-19 and to treat COVID-19. Some potential therapeutic effects of CBD on pain, nausea, and epilepsy are confirmed by NIDA,(34) but there is no evidence to support that CBD protects users against COVID-19. Another video suggested that vaping “bee juice” (propolis) can prevent COVID-19. Propolis may be a healthy substance for strengthening immune systems(35); however, there is no evidence to support that use of these substances or vaping these substances can protect individuals from getting or treating COVID-19. Given that the effect of vaping such substances on COVID-19 is still unclear,(36–40) these videos are disseminating unconfirmed health information.

Notably, a number of “*face mask-related*” videos existed. For example, videos showed comparisons of vaping with different types of cloth, surgical, and N95 masks and vaping while wearing a mask. Some aerosols were retained in masks, which may cause re-inhalation. This is concerning since re-inhalation may increase exposure to nicotine and chemicals from vaping.(41) Further, we found a video instructing how to modify a vaping device for use while wearing a mask. Consistent with prior studies,(12,42) e-cigarette modification videos still persist, but are now related to COVID-19. Taken together, these results show that videos endorsing risky behaviors and have gained attention from YouTube users. Surveillance is warranted to continuously identify videos that might mislead or endanger viewers.

We observed more likes and comments for “*instructional*” and “*sales/access*” videos than for videos with other themes. Even though the comments were not systematically analyzed, it appeared that the comments were positive towards vape retailers and negative towards governmental decisions regarding vape shop closure during COVID-19. For example, the positive comments showed appreciation of entrepreneurship (e.g., “*amazing to see a business going so far for their customers, respect*!”). The negative comments were geared toward governmental decisions regarding vape shop closure during COVID-19 since vape shops were considered “non-essential” businesses (e.g., “*all businesses are essential. Regardless, telling a business they can’t open is unconstitutional. Polis [sic*.*] should be sued and arrested for illegally imposing orders that violate citizens constitutional rights*”).

Video themes in the “*other*” category included an upscale party (e.g., women wearing formal gowns and jewelry, candlelight in the patio party room) in Bali Indonesia where party attendees were vaping. A video in the “*other”* category of a party abroad that took place during the travel ban was uploaded by a vape shop. Such videos are concerning because of the portrayal of vaping as cool and glamorous, especially during the travel ban during COVID-19.

In summary, we identified a variety of e-cigarette-related content during the COVID-19 pandemic, which includes health information related to e-cigarettes and COVID-19, e-cigarette sales and access during COVID-19 (e.g., vape shop closure), and face-mask-related e-cigarette videos. Nonetheless, we acknowledge several limitations. First, we analyzed only videos in English and there may be other content related to COVID-19 not captured in English. Since YouTube is a social media platform used globally and e-cigarette use and COVID-19 are global health issues, future studies would benefit from examining non-English content. Second, we examined e-cigarettes and COVID-19-related content only on YouTube. Future studies should examine other social media platforms as the content may differ. Third, future studies should systematically analyze the sentiments of the comments to better understand perceptions related to misinformtaion related to e-cigarettes and COVID-19 to communicate accurate health information on social media.

Currently, COVID-19 is still widespread globally. As the health impacts regarding e-cigarette use on COVID-19 are less known, future studies should examine the association between them. Future studies should also examine how health information regarding e-cigarettes and COVID-19 is portrayed on social media such as YouTube, and how COVID-19 was used for e-cigarette promotion. Continuous surveillance and the monitoring of novel vaping-related content in reaction to policies and events such as the global pandemic on social media are urgently needed.

## Data Availability

The data underlying the results presented in the study are available from https://www.youtube.com/

## Notes

**FUNDING:** This study was supported by R01DA049878 (PI: G, Kong).

**CONFLICT OF INTEREST:** None

### Competing Interest Statement

The authors have declared no competing interest.

### Funding Statement

FUNDING: This study was supported by R01DA049878 (PI: G, Kong). Funders did not play any role in the study design, data collection and analysis, decision to publish, or preparation of the manuscript.

### Author Declarations

This is an observational study of publicly available social media data.

